# Impact of the COVID-19 pandemic on ongoing health research: an ad hoc survey among investigators in Germany

**DOI:** 10.1101/2020.08.14.20174888

**Authors:** Tanja Bratan, Heike Aichinger, Nicole Brkic, Jana Rueter, Christian Apfelbacher, Julika Loss

## Abstract

**Objectives:** To gain insights into the impact of the COVID-19 pandemic on ongoing health research projects, using projects from a selected funding programme in Germany as an example.

**Design:** Online survey

**Setting:** Lockdowns and social distancing policies impact upon clinical and public health research in various forms, especially if unrelated to COVID-19. Research institutions have reduced onsite activities, data is often collected remotely, and during the height of the crisis, clinical researchers were partially forced to abandon their projects in favour of front-line care and crisis response.

**Participants:** 120 investigators of health research projects across Germany, performed between 15 and 25 May 2020.

**Results:** The response rate (78%) showed that the survey generated significant interest among investigators. 85 responses were included for analysis, and the majority of investigators (93%) reported that their projects were affected by the pandemic, with many (80%) stating that data collection was not possible as planned, and they could not carry out interventions as planned (67%). Other impacts were caused by staff being unavailable, for example through child or elder care commitments or because of COVID-19 quarantine or illness. Investigators also reported that publications were delayed or not feasible at all (56%), and some experienced problems with PhD or Masters theses 18%). The majority of investigators had mitigation strategies in place such as adjustment of data collection methods using digital tools (46%) or of project implementation in general (46%), others made changes in research design or research questions (27%).

**Conclusions:** The COVID-19 pandemic has severely impacted upon health research projects. The main challenge is now to mitigate negative effects and to improve long-term resilience in health research. The pandemic has also acted as a driver of innovation and change, for example by accelerating the use of digital methods.

**Strengths and limitations of this study:**

- To our knowledge, this is the first study investigating the impact of the COVID-19 pandemic on non COVID-19 health research projects, mitigation strategies employed by investigators and needs for support.
- The sample is representative of the projects from the “Healthy - for a lifetime” funding programme in Germany, which includes different types of health research projects and involves different population groups.
- We were not able to clearly distinguish the effects on different types of projects (clinical studies, observational studies, secondary data analyses etc.), because a small number of investigators led more than one project and were not asked to report on each project individually.
- The survey presents a snapshot of the situation in May 2020. To assess effects more widely as well as long-term impacts on projects, the survey would need to be repeated.

## Introduction

Since its outbreak in Wuhan in the People’s Republic of China at the end of 2019, the novel coronavirus (severe acute respiratory syndrome corona virus 2, SARS-CoV-2) has rapidly spread from its origin in the Hubei province to the rest of the world. It causes COVID-19 disease, primarily affecting the respiratory system, with evidence of the effects on other organs and systems also emerging. COVID-19 was declared a pandemic by the World Health Organization (WHO) in March 2020 (1). The virus is spread from person to person through direct contact and droplets (2) Subsequently, governmental responses worldwide have focused on mitigation strategies such as social distancing, travel and movement restrictions, school closures, restricting group and mass gatherings, up to the banning of public transport and lockdown of offices, services and industries (3, 4, 1).

In most countries, these restrictions have disrupted people’s lives and work in an unprecedented way (5, 6). The pandemic has also impacted upon clinical and public health research in various forms. On the one hand, the pandemic has placed scientific virologists, epidemiologists and pneumologists at the forefront of COVID-19 research, and the number of academic publications on COVID-19 is soaring (7). On the other hand, maintaining clinical, health services research and public health studies is considerably impeded by lockdowns and social distancing policies. Many research institutions have severely reduced onsite research (8), research activities have to be performed remotely (especially research unrelated to COVID-19), and during the height of the crisis clinical research programs were forced to abandon their schedules in favour of front-line care and crisis response (9). Personal contacts with study participants and meetings among research partners needed to be cancelled (8) or restricted.

This is also a set-back for public health and health services research. The strengthening of diversity aspects as well as patient and civil rights over the past decades has transformed health-related research: patient and public involvement in the planning and evaluation of clinical studies and in health promotion have evolved to be the gold standard (10). Studies now prefer ‘real life’, complex interventions engaging multiple stakeholders and partners in settings and health care institutions (11). In order to assess the effectiveness of these multi-level interventions, mixed-method designs have become increasingly popular, as they combine standardised measurements and surveys with intensive qualitative data collection methods such as interviews and focus group discussions (12-14).

These achievements in health-related research may have now made this kind of research particularly vulnerable to social distancing measures and stay-at-home policies. Settings such as nursing homes or schools cannot be approached easily anymore, participatory in-person meetings with stakeholders, patients or citizens are not possible or made difficult, as are face-to-face data collection methods. Inouye et al., report that field researchers may have to abandon an entire field season due to bans on traveling and recruiting, and thereby lose irreplaceable data (15).

In addition, parents face novel challenges induced by closures of schools and day care centres, as they need to devote time to looking after and home schooling their children and doing household chores. Combining child care needs with remote academic working can prove difficult, if not impossible in many cases (16, 17). This may further slow health-related research.

Few editorials and opinion pieces have raised awareness for the potentially substantial constraints that the COVID-19 pandemic places upon the efficiency of ongoing scientific proceedings (8, 16, 15, 9) but empirical studies exploring or quantifying the challenges and needs of researchers engaged in ongoing health research unrelated to COVID-19 are lacking to date.

Therefore, we intended to understand

- if, and how, non-COVID-19 related health research is affected by the COVID-19 pandemic,
- what strategies are used by researchers to mitigate challenges and potential (academic) damages to their projects.

We addressed these questions by surveying investigators who are responsible for research funded by the funding programme “Healthy - for a lifetime”. This is a four year governmental funding programme in Germany (2017 - 2021) with an emphasis on the development and evaluation of new concepts for health promotion, prevention and care for different life phases.

## Methods

### The funding programme “Healthy - for a lifetime”

In Germany, the Federal Ministry of Education and Research is, apart from the German Research Foundation, the main funding agency for research (18). In health, main funding activities relate to preventing and tackling common diseases, health services research, prevention and nutrition research and personalised medicine (19). In 2016, the Ministry launched the ‘Healthy - for a lifetime’ funding programme (*‘Gesundein Leben lang’)* to better address the following groups: children and young people, the working population, older people as well as men and women. For the research initiative, the Federal Ministry has provided approximately 100 million euros in funding to promote the development of new and effective concepts for health promotion, prevention and care. In total, 174 single projects and subprojects as part of consortia are being funded in 79 different German universities or research institutions. The funding programme consists of projects in five funding areas: Gender health (n = 32), occupational health (n = 35), child and youth health (n = 60), clinical studies in old age (n = 18) as well as healthcare and nursing studies in old age (n = 29). The majority of these projects can be defined as health services research or prevention research in the form of interventions (53%); fewer studies relate to literature reviews and studies with existing data (20%), observational studies (17%) or biomedical/ laboratory research (3%).

The survey presented in this study is part of an accompanying research project for the ‘Healthy - for a lifetime’ initiative (GeLang-Bella^1^), the aim of which is to establish networks between projects of the funding programme, offer scientific support, and to develop standards for central overarching themes such as participatory approaches, patient-related outcomes, or transfer of research results to practice. Its advisory board includes several patient representatives.

### Participants

We performed an ad-hoc single online survey among researchers responsible for projects funded within the funding programme ‘Healthy - for a lifetime’. All investigators who had agreed to participate in the accompanying research project GeLang-BeLLa (N = 120) were sent an e-mail invitation with a personalised link to the survey. Investigators who were in charge of more than one project (n = 10) were only sent one link, and for their convenience were asked to jointly consider all of their projects in their response.

### The online survey

The survey was implemented as an online version using EFS Questback and was available between 15th and 29th May 2020. The email invitation to complete the survey was followed up by two reminders. A multi-option structured response format was used. In addition, free text fields were provided to allow participants to add individual comments. The survey consisted of five items enquiring (1) How the pandemic impacted on project implementation, process, and results, (2) which specific (organisational, personal,…) conditions had caused this impact, (3) whether academic output was compromised, i.e. concerning publications or master’s or doctoral theses, (4) which type of mitigation strategies had been implemented, and (5) whether there was a need for specific support measures from the accompanying research project.

### Statistical analysis

Data generated were analysed descriptively using Microsoft Excel. All variables were categorical, hence counts and percentages were computed.

### Ethical considerations

All 144 principal investigators of the 174 studies (some lead two or more studies, see above) were asked to give informed consent for data collection and data storage for the accompanying research project, including the consent to (a) be sent an online questionnaire, to (b) have the questionnaire data analysed and saved. 120 principal investigators gave their written consent and were included in the study. All questionnaires were de-identified by an independent trust centre before analysis. The study was approved by the Ethics Committee of the University of Regensburg (19-1630-101).

### The online workshop

A one-hour online workshop open to all interested investigators from the ‘Healthy - for a lifetime’ funding programme was held on 28 May 2020. Thirty-two investigators participated in the virtual event. They were presented with the results from the survey and asked to discuss them. The workshop was minuted, and the minutes were analysed with regard to (a) confirmation of presented study results, and (b) additional aspects that were brought up in response to the research questions, given changes in COVID-19 mitigation policies that have emerged within the timespan after the survey.

## Results

### Sample

Out of the 120 investigators who were invited to participate, 93 (78%) completed the questionnaire. 8 responses were excluded from the sample because the projects had already ended and could therefore not have been affected by the pandemic, which led to sample of 85 (71%) questionnaires for analysis.

All funding areas are represented in the survey, with child and youth health projects most prevalent. The distribution across the different funding areas broadly matches the overall distribution of all funded projects, with gender projects and clinical studies in old age being slightly underrepresented in our sample and healthcare and nursing studies in old age being very slightly overrepresented. A small number of respondents were unsure which funding area their project could be assigned to.

### Impact of the COVID-19 pandemic on research

The vast majority of investigators reported that their projects were at least partially affected by the pandemic, either because implementation was being impeded through the crisis (84%), or because it was suspended (18%) (Figure 1).

**Figure 1:**
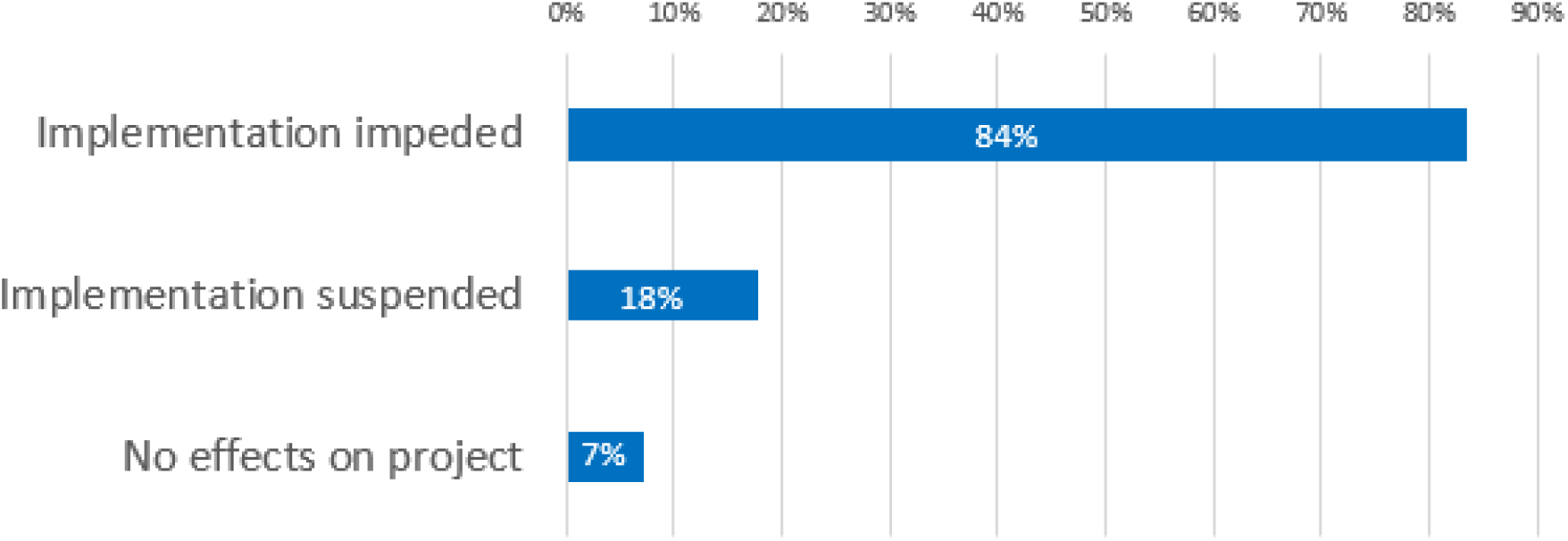
Perceived effects of the COVID-19 pandemic on project implementation, N=85, multiple answers possible

Those respondents who reported an effect on project implementation (93%), were asked for the causes (Figure 2). The most frequently cited barriers to continuing research projects were difficulties in data collection procedures (80%), and failure to implement planned interventions (67%). Also, staff shortages due to the pandemic were reported, e.g. due to child care commitments during the lockdown resulting from the closure of child care facilities or because of elder care commitments (38%), due to COVID-19 quarantine and disease (11%) or because all project work had been suspended because of official instructions (14%) or because staff had been assigned to other tasks, e.g. clinical work (9%).

**Figure 2:**
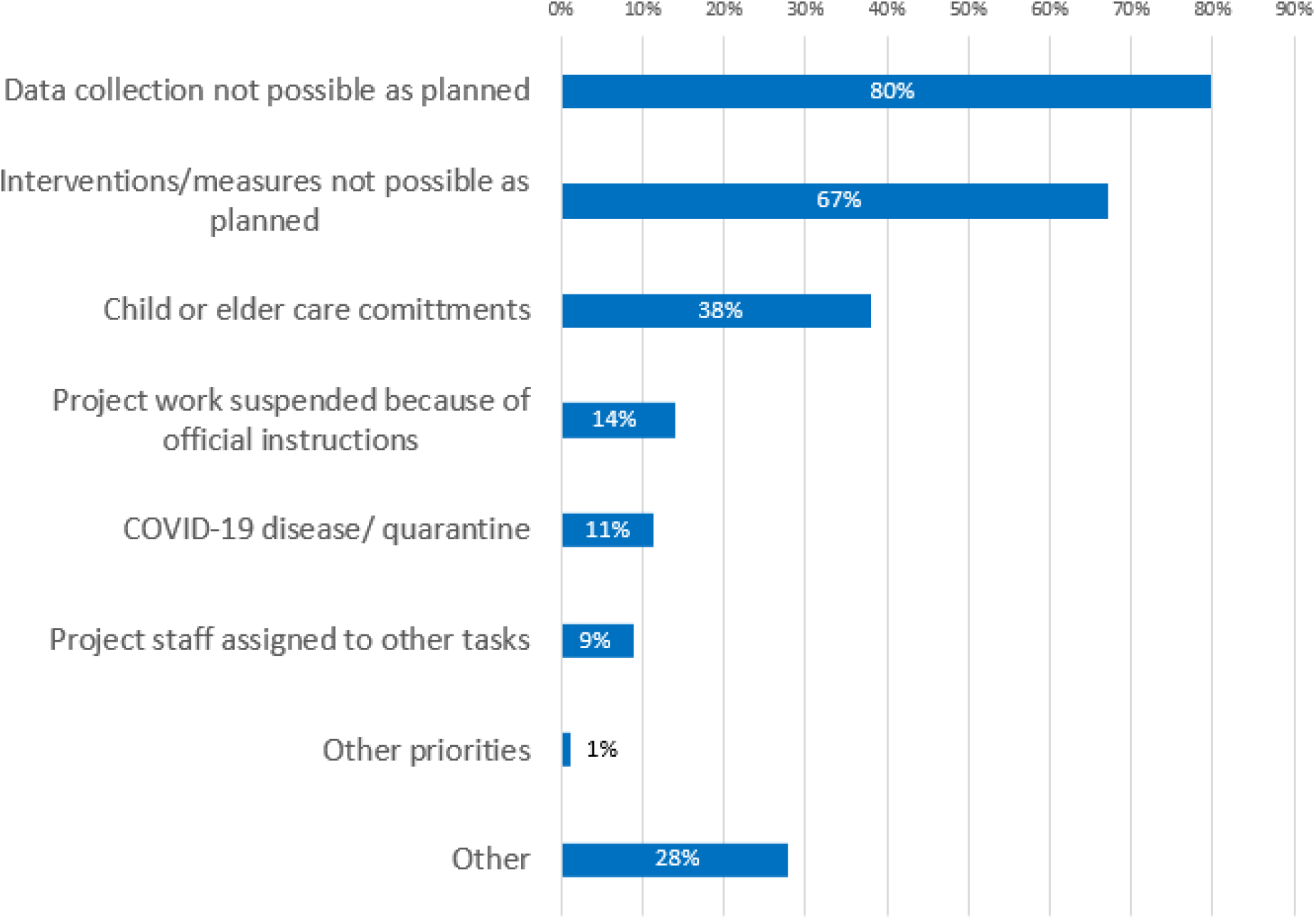
Causes of research impediments, N=79, multiple answers possible

**Figure 3:**
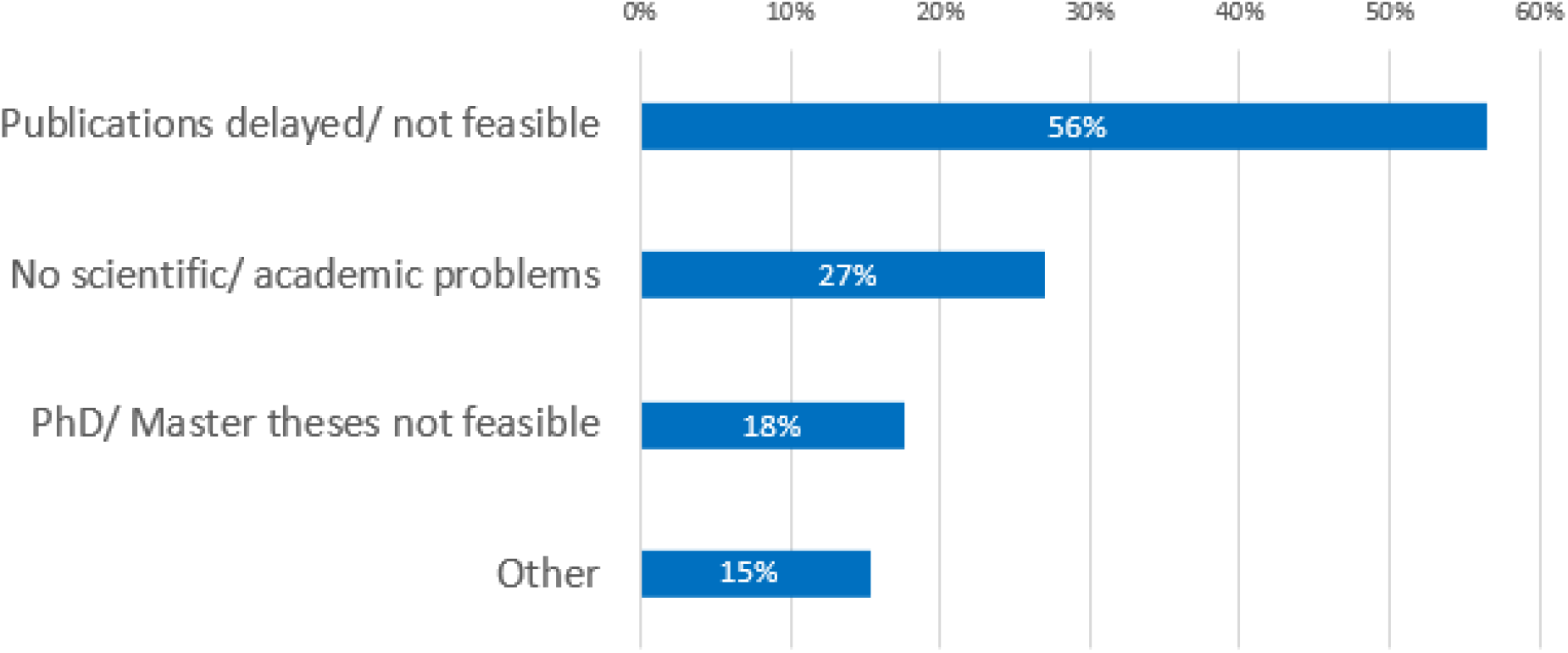
Influence of the pandemic on scientific and/or academic progress, N=85, multiple answers possible

Additional free text responses indicated further problems with recruitment of study participants, which proved more difficult during the pandemic, had been put on hold or ended ahead of time (n = 4). Data collection was described as being more difficult or of lower quality (n=3). Practical adjustments such as shifting tasks between project partners, working from home and virtual meetings replacing travel were also reported (n=6), while others cited difficulties caused by working from home, which included access to data or technical infrastructures (n=2). Lacking possibilities of validating findings through conference presentations were also mentioned (n=1) The COVID 19 pandemic also impacted on scientific outputs and academic careers; for example, more than half of participants stated that publications were delayed or could not be realised (56%). Difficulties with continuing PhD and master’s theses were also reported (18%). Figure 6 shows more details.

The majority of researchers have reacted to the restrictions caused by the pandemic with mitigation strategies. They modified their data collection methods (46%, e.g. by employing Internet-based access to study participants) or made adjustments in project implementation (46%). In some projects, the research concept including the research questions were adjusted, sometimes to include COVID-19 related topics (27%). However, some projects (18%) did not employ mitigation strategies, sometimes because no suitable measures were available. See Figure 4 for more details.

**Figure 4:**
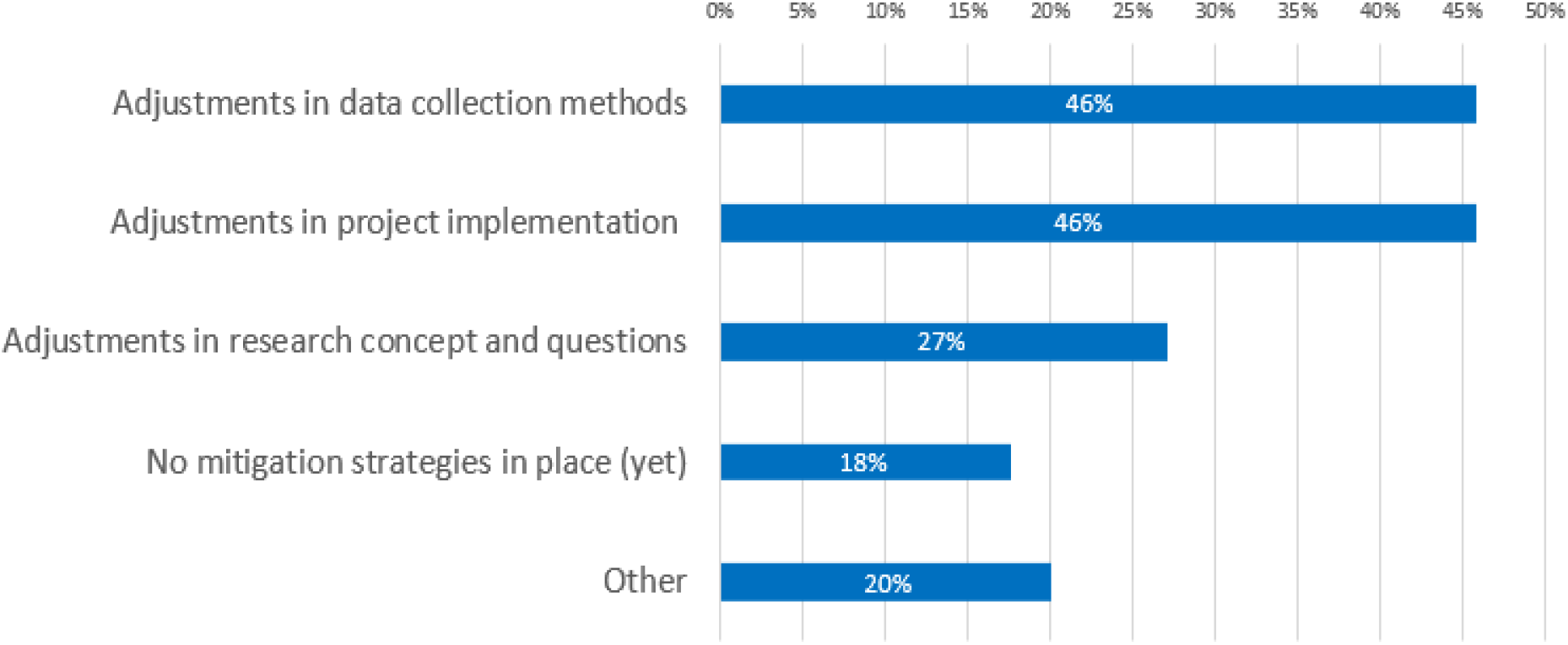
Mitigation strategies used to deal with restrictions caused by the pandemic, N=85, multiple answers possible

In terms of support requirements, some researchers expressed an interest in sharing know-how with the other funded projects about digital communication tools (21%) and enabling participation digitally, e.g. in terms of organisational and moderation skills (19%).

**Figure 5:**
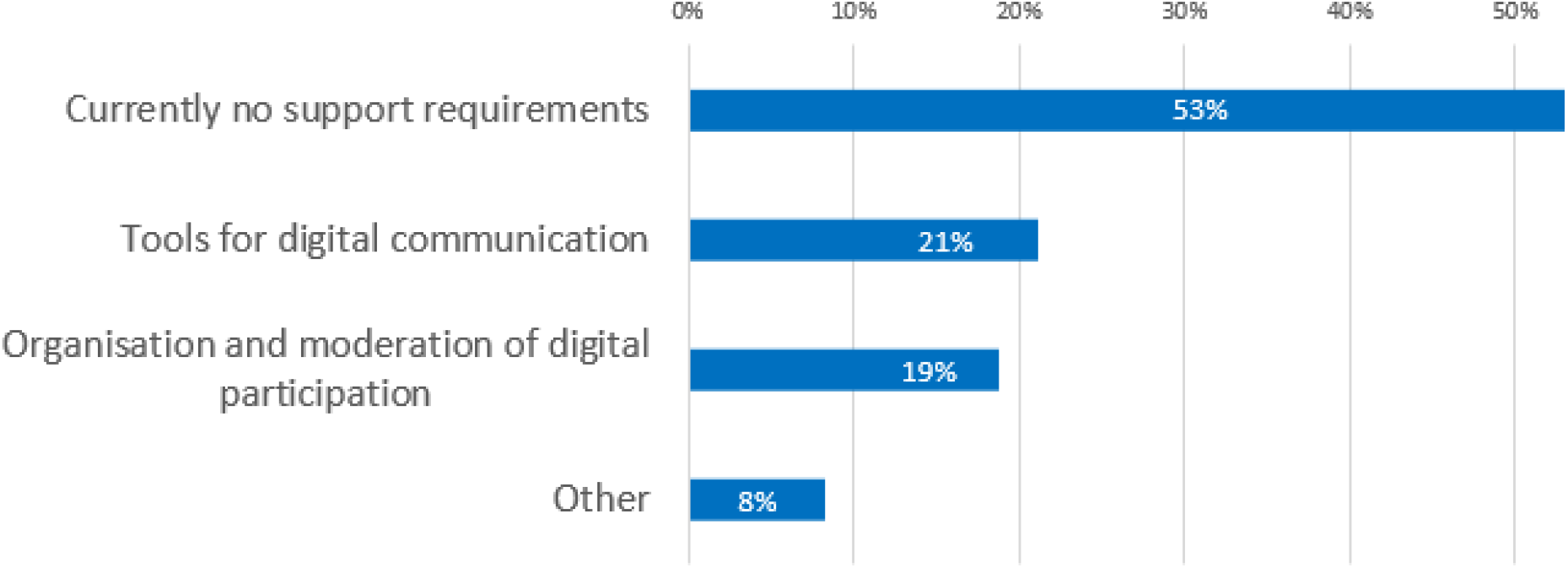
Need for support from the accompanying research project

### Validation through online workshop

The discussions documented during the workshop confirmed the survey results and highlighted the immense effect the pandemic has had on many health research projects not related to COVID-19. Concerns were raised about the ability to re-start interventions, for example in the case of workplace interventions when staff were working remotely or were on reduced hours.

Investigators of projects that were able to continue implementation were concerned about the validity of their research in the face of deviations from study protocols that had been necessary during the pandemic. It was also pointed out that some projects, e.g. about mental health, had defined patient endpoints such as loneliness and depression, which were now severely affected by the pandemic, so the comparability of the data to earlier results may be reduced.

Organisational issues regarding data collection and implementation of interventions during the pandemic were raised, e.g. the need to inform participants about risk of infection, hygiene requirements or liability. Difficulties of elderly participants with online data collection were reported as a further practical challenge.

## Discussion

### Principal findings

Our findings show that the COVID-19 pandemic has had a severe impact on the vast majority of 93 heterogeneous health research projects of the “Healthy - for a lifetime” funding programme. The programme is not related to COVID-19 research, and most projects were unable to continue their work as planned. They were impeded in their recruitment of participants, implementation of interventions or data collection. A lack of staff availability due to private or other professional commitments or as a result of COVID-19 quarantine or illness were also observed by half of the investigators surveyed. Several participants reported that projects had to be suspended temporarily, and at the time of the survey, it was not clear whether they could be resumed in the near future. Investigators were creative in developing mitigation strategies for restrictions in data collections, with many drawing on digital communication, but this was not an option for all projects. A quarter of participants stated that they bridged the imposed suspension in their projects by adjusting their research, including pursuing novel COVID-19-related research. Investigators also expressed a need for exchange on digital communication as well as guidance regarding issues such as hygiene and participation. Methodological issues related to deviation from study protocols or validity of mid-study changes in data collection methods. This also raised concerns as to whether the data would eventually qualify for publication and further scientific exploitation, or whether they would ultimately need to be abandoned.

### Meaning of study and implications for policy and practice

Due to the ongoing need for social distancing, personal contact with study participants and therefore resumption of regular data collection and implementation of interventions is likely to remain difficult. Also, there is the risk of new waves of infections and either local or general lock-downs due to SARS-CoV-2, but possibly in the future also due to other pandemics. Therefore, future strategies for planning, implementing and funding health research need to incorporate the possibility of potential disruptions and restrictions inflicted by pandemics and infection control measures. The importance of research, especially in crisis situations, as well as the need for new paradigms and models of resilient and efficient research has been highlighted in the literature (20, 21). Therefore, it seems important to not only handle the current challenges, but also to plan for long-term approaches preventing or taking into consideration these challenges for future research. Our study raises the following important questions: a) How can progress made with participation in health research be maintained despite difficulties and uncertainties about the future? b) How can resilience be built into study protocols to ensure that they can be adapted if necessary and data already collected is not lost, and at the same time protocols remain methodologically robust? c) How can different intervention and data collection methods be meaningfully combined and biases introduced be accounted for? d) How can funding instruments be designed to accommodate changes more easily and, e) How can funders support investigators during crises such as pandemics?

The pandemic is currently changing the way scientific knowledge is being produced (22), in fact it is accelerating a trend that has already been underway: The use of digital tools had been increasing gradually (23, 24), and during the pandemic, with often no other alternatives being available, it has surged (25). While the pandemic undeniably poses many challenges to health research projects, its silver lining may be a chance to make a leap in digital communication and participation as well as better resilience at both the research and the funding side. This should be accompanied by a thorough investigation of the strengths and weaknesses as well as the comparability of different tools for interventions and data collection methods. Existing findings on the comparison of analogue and digital data collection methods are sparse and limited in scope, but so far indicate that there are no far-reaching differences (26–31). More research is required regarding issues such as acceptance, reach and over-/ underrepresentation of different groups, usability in different settings and for different topics. During data analysis, the influence of changes in collection methods and other deviations from study protocols as well as missing information need to be considered. Descriptions should delineate which of these irregularities are likely to be a result of the COVID-19 pandemic and which uncertainties remain (32).

Funders should consider granting extensions to projects if these face delays because of the pandemic, and allow for adjustments in research design and research questions. Changes to funding itself may also be needed. To prepare for ongoing restrictions, further lockdowns or other pandemics, policy makers and funders could introduce more flexible funding instruments. Research that generates evidence about the validity and scientific rigour of digital methods or about the combination of digital and traditional methods will be needed to accompany the shift to a “new normality” in research, and it will also be a task for policy makers to ensure research priorities are set accordingly.

Researchers will be required to continue experimenting with new approaches, assessing their usefulness, reflecting on their findings and sharing their insights. The scientific community at large will have to deal with results of research that have taken place under different circumstances than usual, and maybe with methodologic compromises. Consensus will be needed about how these findings can be meaningfully integrated with other scientific outcomes, both in terms of comparison to existing findings and in terms of research validation. Ultimately, it is a joint responsibility of policy makers, researchers, health professionals and funders to ensure that research funding is spent efficiently and effectively. The pandemic has changed the way in which scientific knowledge is produced, and some of the changes may be permanent, which will ultimately require adaptions to what constitutes good scientific practice.

### Strengths and weaknesses

To our knowledge, our study is the first ever investigation into the effects of the pandemic on health research projects not related to COVID-19. It uses a representative sample of heterogeneous projects from the “Healthy - for a lifetime” funding programme in Germany, therefore giving important insights into the impact on health research in general. However, it only presents a snapshot of the situation in May 2020, and captured the experiences of a limited number of investigators. Due to the dynamics in the COVID-19 pandemic and infection control measures, restrictions in project work and data collection processes vary significantly over time. Therefore, it would be helpful to repeat the survey at certain intervals.

The research projects included in the study covered a wide range of topics, addressed target groups, and study types. However, due to the overall focus of the funding programme, there was a predominance of intervention projects in prevention and health services research; biomedical research accounted for only a minority of projects. Among biomedical, laboratory-based studies, regulations about social distancing may have a different influence (e.g. by rendering access to labs difficult, rather than preventing contact to patients or participants). Still, our survey results highlight the range and extent of challenges imposed upon health research.

## Conclusions

The disruption of health research projects caused by the COVID-19 pandemic has been severe and calls for short-term measures to limit damage to projects and to participation in health research in general, but also the development of long-term strategies to improve the resilience of research against imponderables posed by pandemics. Both require flexibility from policy makers, funders and researchers as well as insights and guidance from the scientific community.

What is already known on this topic

- To our knowledge, there has been no previous research on the impact of the COVID-19 pandemic on ongoing health research projects.

What this study adds

- The study sheds light on how ongoing health research projects in Germany have been affected by the COVID-19 pandemic. It also investigates what mitigation strategies have been put in place, what issues could not be resolved and what challenges and opportunities the current situation holds for policy makers, funders, researchers and the scientific community at large.

### Footnotes

#### Contributors

TB, JL, CA, HA and JR designed the study. TB led its implementation and together with JL wrote the initial draft of the article. All authors were involved in subsequent protocol revisions. NB carried out the initial analysis of the data and all authors were involved in further analysis.

#### Ethical approval

University of Regensburg Ethical Committee (19-1630-101).

#### Data sharing statement

No additional data are available.

TB affirms that the manuscript is an honest, accurate, and transparent account of the study being reported; that no important aspects of the study have been omitted; and that any discrepancies from the study as planned (and, if relevant, registered) have been explained.

Dissemination to participants and related patient and public communities: Preliminary findings have already been shared and discussed with investigators from the “Healthy - for a lifetime” funding programme. The publication will also be disseminated among this group, the funding body and beyond.

A preprint of this manuscript has been deposited at https://www.medrxiv.org/.

## Data Availability

All data generated or analysed during this study are included in this published article (and its supplementary information files).

## Footnotes

1 Project website https://www.begleitforschung-bella.de/

